# Unmasking Seasonal Cycles in Human Fertility: How holiday sex and fertility cycles shape birth seasonality

**DOI:** 10.1101/2020.11.19.20235010

**Authors:** L. Symul, P. Hsieh, A. Shea, CRC. Moreno, D.J. Skene, S. Holmes, M Martinez

## Abstract

The mechanisms of human birth seasonality have been debated for over 150 years^1^. In particular, the question of whether sexual activity or fertility variations drive birth seasonality has remained open and challenging to test without large-scale data on sexual activity ^2,3^. Analyzing data from half-a-million users worldwide collected from the female health tracking app Clue combined with birth records, we inferred that birth seasonality is primarily driven by seasonal fertility, yet increased sexual activity around holidays explains minor peaks in the birth curve. Our data came from locations in the Northern Hemisphere (UK, US, and France) and the Southern Hemisphere (Brazil). We found that fertility peaks between the autumn equinox and winter solstice in the Northern Hemisphere locations and shortly following the winter solstice in the Southern Hemisphere locations.

## Main Text

To our knowledge, the earliest identified written report of human birth seasonality was nearly 200 years ago^1^. Since then, a large body of scientific literature has grown around the topic. A consistent and striking pattern is that Northern Hemisphere countries—where most work on birth seasonality has focused—had much stronger birth seasonality in the pre-industrial and early industrial era, compared to dampened seasonality today^2–4^. This dampening is likely driven by multiple factors that diminished human connectedness to the natural environment, which we discuss below, and/or behavioral practices that have been culturally modified over time. It remains unclear if modern-day birth seasonality emerges from changes in sexual activity or seasonal fertility, broadly defined here as the ability to produce offspring.

Evidence for seasonal sexual activity has been presented in studies from multiple countries that have found correlations between holidays, including fertility festivals, and the timing of peak conception ^5,6^. Thus, the dampening of birth seasonality over time could be due to cultural changes that have made sexual activity less seasonally structured. Similarly, improved family planning and birth control development may have resulted in births becoming more uniformly distributed throughout the year ^7^ or distributed around seasons most desired by couples ^8–10^. In addition to seasonal sexual activity, biological factors that may play a role in birth seasonality include (i) seasonal biological rhythms in human fertility entrained by environmental light intensity/photoperiod^11,12^, (ii) seasonal variation in male fertility driven by temperature effects on sperm^13^, (iii) seasonal variation in female fertility driven by the seasonal availability of fat-rich food^14,15^, and (iv) seasonal variation in early pregnancy loss due to environmental hardships such as seasonal malnutrition or intense physical labor^16,17^. All hypothesized mechanisms may be expected to dampen as societies become less subject to the natural (seasonal) environment.

From a public health perspective, birth seasonality has three potential implications. First, it has direct consequences on maternal and infant health. Each country has a gestational peak, a birth peak, and a nursing peak. Any one of these peaks may coincide with environmental challenges – such as seasonal disease outbreaks, elevated food insecurity, and extreme weather events (e.g., heat waves, cold spells, flooding, etc.) – and can put more pregnant individuals and/or newborns at risk. Second, birth seasonality impacts sexual and reproductive health; health services relating to contraception, miscarriages, abortion, and/or sexually transmitted infections (STI) may be needed more at particular times of the year. Seasonal changes in sexual activity may increase the need for contraception and family planning services at specific times of the year. On the other hand, a high fertility season may pose an unrecognized time of risk for unwanted pregnancy, which can be a danger for women and girls in many countries. Lastly, if birth seasonality is driven by seasonal variation in fertility, it could inform fertility treatments, especially for those involving the endogenous hypothalamic-pituitary-ovarian (HPO) axis in females; this includes ovulation induction and intrauterine insemination. To date, there is conflicting evidence regarding seasonal effects on fertility treatments and assisted reproductive technologies (ARTs) ^18–23^. There remains a need for a critical understanding of the drivers of birth seasonality and whether human fertility is seasonal; this knowledge may be invaluable for improving reproductive health.

The primary objective of this study was to quantify the contribution of sexual activity *vs*. fertility to seasonal variations in births. While population-level birth data are accessible through governmental vital statistics, data quantifying sexual activity and fertility are challenging to obtain. Here we used a digital epidemiology approach applied to high-resolution data of sexual activity from half a million users of a menstrual cycle tracking app, Clue, available worldwide. Clue is used by individuals of various ages, reproductive objectives (pregnancy, contraception, or not applicable), sexual orientation, gender identity, and cultures. On a daily basis, Clue users can log their menstrual symptoms (*e*.*g*., bleeding, pain, etc.), sexual activity, and birth control use. These individual-level data were then aggregated to construct population-level time series of relative sexual activity throughout the year. We then established mathematical models to simulate births under multiple scenarios while making the following assumptions: (i) births follow conceptions by a delay estimated from empirical distributions of gestation times, (ii) conceptions are modeled as the product of sexual activity and fertility, and (iii) fertility is assumed to be a composite term including male and female fertility at conception as well as the rate at which pregnancies are carried to term (Methods). We tested three scenarios to determine whether seasonal sexual activity and/or seasonal fertility shape birth seasonality by fitting the models to data from six populations and comparing the likelihood of these three scenarios. We also provided descriptive statistics of sexual activity in the US, France, UK, and Brazil, investigated the impact of age, birth control methods, and condom use on our inference, and estimated peak timing and amplitude for seasonal fertility.

### Birth Seasonality

Our study focused on six geographically and culturally diverse locations: U.K., France, Central-West Brazil, Northeast Brazil, California, and the Northeastern US (Fig 1a). We collated time series data of monthly births for each location (Methods). Data from all six locations displayed birth seasonality with variation in amplitude and peak timing from country to country (Fig 1a). The birth peak timing ranged from March-May in Brazil for the major birth pulse, while September displayed a minor/smaller birth pulse. Births peaked from July-Sept in France and the Northeastern US, while the UK and California had their peak in September.

**Fig 1.**
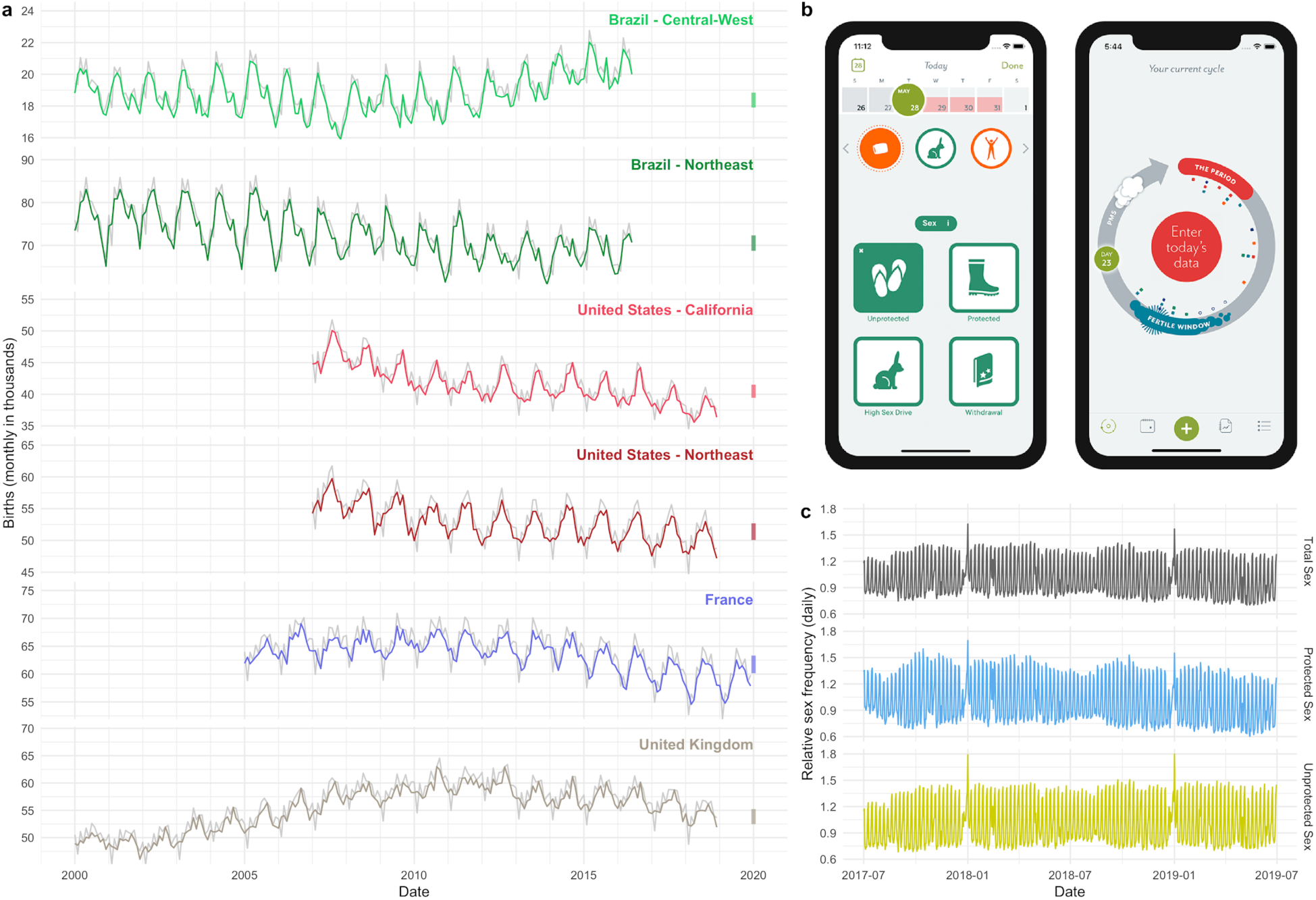
Birth seasonality and sexual activity. (a) Time series of monthly births for each location in the study. Gray lines are the raw data, overlaid by births standardized to a 30-day month (in color). Vertical bars on the right axis represent a 5% amplitude relative to the mean, for reference. (b) Snapshot of the Clue app smartphone interface. (c) Daily relative sexual frequency aggregated for all locations and stratified by sex type: protected, unprotected, and total sex. These time series are available for each location and are also shown aggregated monthly in the Supplementary Material (Suppl. Fig. 1, 27).

### Sexual Activity

We used sexual activity data collected from over 500,000 individuals in our six study locations, representing 180 million days of active tracking data. The data were provided by Clue by BioWink

GmbH^24^, a women’s health mobile phone app (Fig 1b). We aggregated the individual-level sexual activity data into population-level time series of relative sexual activity. We observed patterns of sexual activity, specifically spikes on weekends and holidays (Fig 1c).

To infer how relative sexual activity is impacted by holidays, weekends, and seasons, we used a statistical model to express sexual activity as a combination of day-of-week, month-of-year, and whether it was a holiday. The dominant axis of variation in sexual activity was the day-of-week, with all countries having elevated sexual activity on weekends and decreased sexual activity on weekdays (Fig 1c, Supp Fig 1, 18-26). Our analysis revealed that all locations in our study had elevated sexual activity on holidays (Fig 2, Suppl Fig 1, 2, 31-36). Typically, each country had elevated sexual activity 1-3 days before and after each holiday. For all locations, the time between Christmas and New Year had the highest level of sexual activity.

**Fig 2.**
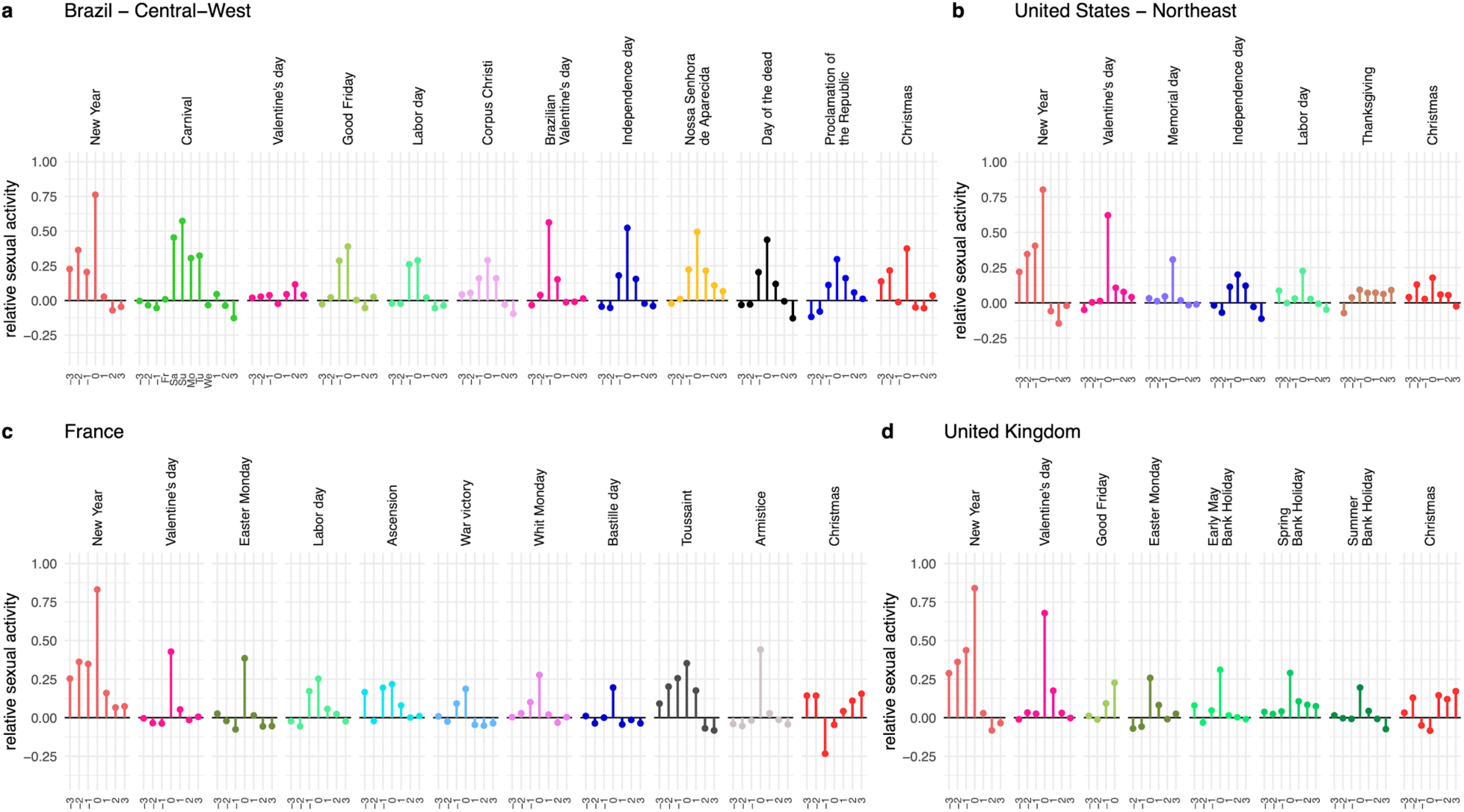
Holiday sexual activity. Relative level of sexual activity for each holiday and each country measured proportional to their mean. For each holiday, 0 on the x-axis refers to the holiday day itself (e.g., Dec 25th for Christmas). Negative values indicate days before the holiday, and positive values refer to the days after the holiday.

To test for biases based on Clue user demographics and behavior, we repeated our analysis by subsetting our data based on (i) age, (ii) birth control method (barrier methods *vs*. hormonal or IUD), and (iii) protected *vs*. unprotected sex. Regarding the age of Clue users, there was not a substantial difference in sexual activity based on age, but we did observe some differences. For example, the difference between relative sexual activity on weekdays vs. weekends was most pronounced for older users in the UK and US, but in Brazil and France, the most pronounced difference was in younger users (Suppl Fig 37). In addition, younger users had more elevated sexual activity on Valentine’s Day than older users. In contrast, at Christmas or Thanksgiving, younger users had lower relative sexual activity than older users (Suppl Fig 40). The birth control method did not have a noticeable impact on sexual activity patterns (Suppl Fig 38, 41). For protected *vs*. unprotected sex, the reporting differed only on select holidays. For most holidays, unprotected and protected sex were equally more frequent; however, around Christmas and New Year, unprotected sex was more frequent than protected sex (Suppl Fig 42).

## Mathematical Model Results

We explicitly tested three hypotheses to determine whether birth seasonality is driven by seasonal fertility vs. seasonal sexual activity. Hypothesis A: birth seasonality is solely driven by seasonal changes in sexual activity. Hypothesis B: birth seasonality is solely driven by seasonal fertility, and Hypothesis C: birth seasonality is driven by a combination of seasonal sexual activity and seasonal fertility. We translated each of the three hypotheses into a deterministic population-level mathematical model of conception and birth (Eq 12 in Suppl. Material, section “Birth Models”).

In each model, daily conceptions were calculated as the product of daily sexual activity and daily fertility. Model A assumed variation in sexual activity, Model B assumed seasonal variation in fertility, and Model C assumed variation in both (see Fig 3a and Methods section on Mathematical Models). Fertility was a composite term that encapsulated multiple biological processes that could generate seasonal fertility. This includes both male and female fertility at conception (e.g., gamete quality, cervical mucus quality, etc.), the likelihood of implantation, and pregnancy loss. In Models A and C, temporal variation in sexual activity was predicted by the statistical model described above using the Clue data for unprotected sex reported by all users in a given location.

**Fig 3.**
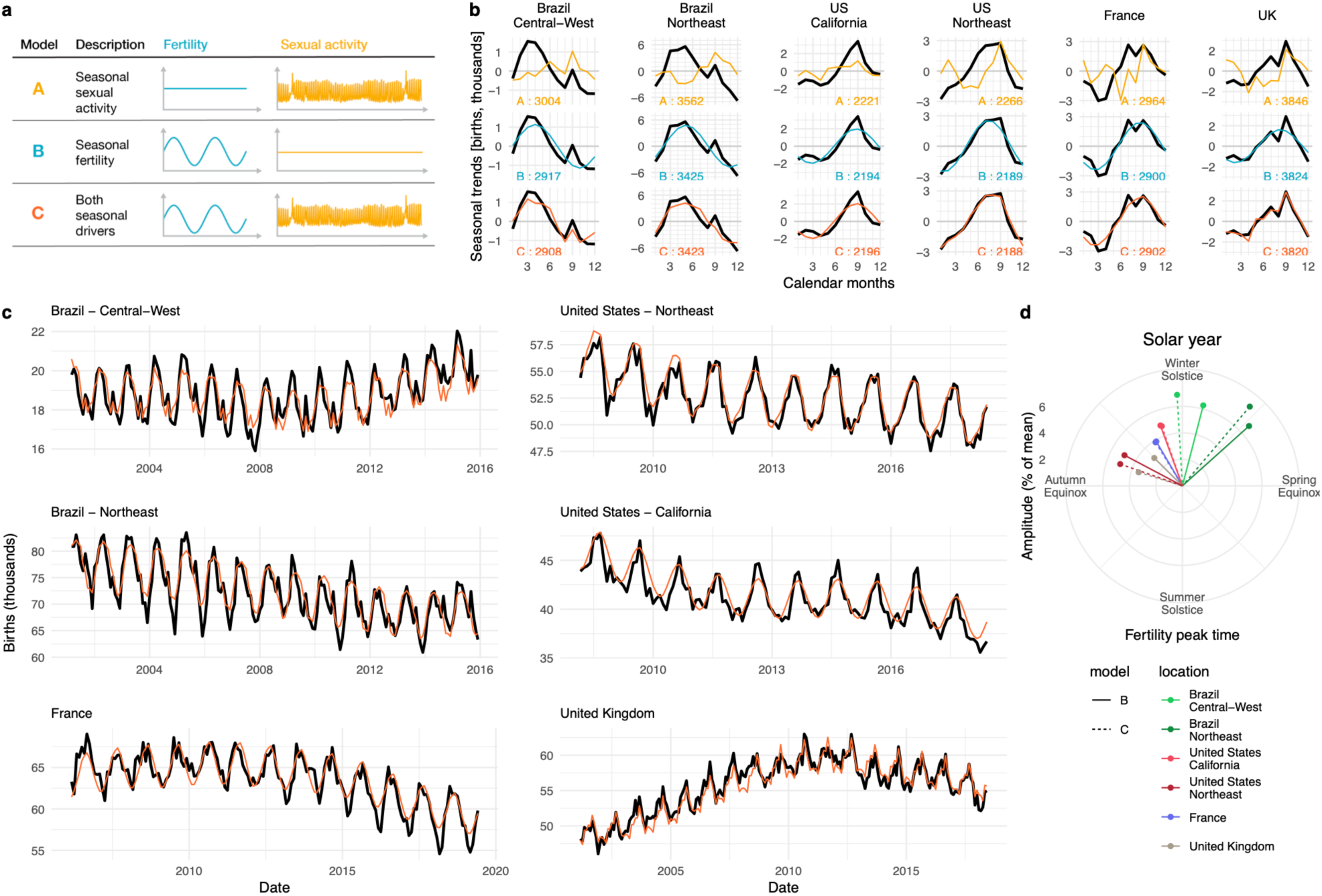
Model results. (a) Schematics of the three variants of our birth model. (b) Seasonal component of time series decomposition for birth data (thick black line) and simulations (thin colored line) from our fitted models (top row: Model A, middle: Model B, bottom: Model C) for each location. Numbers at the bottom of each panel indicate AIC values. (c) Birth time series (thick black line) and Model C simulations (thin colored line) for each location. (d) Timing and amplitude of the fertility peak estimated from Models B and C for each location plotted relative to the solar year.

Daily births were simulated by shifting daily conceptions by a lag corresponding to empirical distributions of gestation periods for each country (approx. nine months into the future) (Suppl. Fig. 17)^25–27^. Simulated births were then aggregated monthly to compare the model outputs with official birth records.

Universally, the models that best fit the data included seasonal fertility (Models B and C). Importantly, Model A, which included seasonal sexual activity as the sole driver of birth seasonality, was insufficient to capture the shape, timing, or amplitude of birth seasonality (Fig 3b top panels, Suppl Fig 3, 54-59, 70-75). To ensure our results were robust to Clue user demographics and behavior, we fit 6-27 model variants using subsets of Clue users for each location. The number of model variants tested depended on sample size within each demographic and behavioral group. We confirmed that Model A is never the best fit. Likelihoods and AICs for all model variants are reported in the Supplementary Materials (Suppl Fig 64-69).

Whether Model B or Model C best fit the data depended on geography. For the UK, Northeastern US, and both locations in Brazil, Model C best fit the data according to AIC, suggesting that both seasonal fertility and holiday sexual activity shape birth seasonality (Fig 3). For France and California, Model B best fit the data, suggesting that seasonal fertility is the dominant driver of birth seasonality in these locations and that seasonal sexual activity plays a negligible role (Fig 3, Suppl Fig 4, Supp Table 10-11 for AIC values and model comparison).

To evaluate the robustness of these results to age-stratification, we re-ran the models using three age categories: ≤23 yr old, ≥24, and all ages. In general, age-stratification did not impact our results or conclusions. The best fit model was the same regardless of age-stratification for five of the six locations; we only found a difference in the Northeastern US where Model B was the best fit for <= 23 yr. old, while Model C was selected for >= 24 and for all ages.

In all study locations, seasonal fertility was supported by the best fit models. To compare seasonal fertility across locations, we characterize seasonal fertility relative to the seasons (solstices and equinoxes) since some of our locations were in the Northern Hemisphere, while others were in the Southern Hemisphere. All locations had their fertility peak between Autumn Equinox and Spring Equinox, centered around the winter solstice. Specifically, Brazil had a fertility peak in Aug-Sept, directly following the winter solstice. Northern Hemisphere locations had their fertility peak from Oct-Dec, directly preceding the winter solstice (Fig 3d). Despite all six locations being located at different latitudes, with different climates and seasons (i.e., temperate seasons at high latitude vs. rainy/dry seasons near the equator), it was remarkable that seasonal fertility peaked within a 3–4-month band centered about the winter solstice.

## Discussion

This study presents evidence that seasonal fertility (as opposed to seasonal sexual activity) is the primary driver of birth seasonality, which has been disputed for 150+ years. While sexual activity variations also contributed to the seasonal birth patterns in some of our study locations, our analyses showed that seasonal sexual activity alone could not explain birth seasonality. Models of birth seasonality require seasonal fertility to capture the data. These results confidently suggest that seasonal reproductive biology exists in humans. We were able to unmask this seasonal phenomenon with our simple mathematical framework because the modern revolution in digital health tracking afforded us the size and resolution of data needed to infer population-level patterns of sexual activity and tease apart the contribution of sexual activity *vs*. seasonal fertility in generating birth seasonality globally.

Past studies of rural and/or nomadic populations that experience dramatic seasonal changes in their living conditions have found mechanisms for seasonal fertility. For example, extreme seasonal changes in birth rates in a nomadic Kenyan tribe were explained by variations in the seasonal availability of fat-rich food and women’s physical labor^28^. In the Lese women of Central Africa, months of low food availability were correlated with reduced salivary progesterone, which may be associated with lower implantation and/or ovulation rates^29,30^. Bolivian women living in high-altitude rural areas, subject to significant seasonal changes in workload, have exhibited decreased salivary progesterone and elevated early pregnancy loss in the months of intensive physical labor^16^. These mechanisms, however important for fertility and successful pregnancy in rural and nomadic populations, may not apply to birth seasonality in industrial or post-industrial countries in which fat-rich food is available throughout the year, seasonal migration is limited, and physical work may be more stable throughout the year.

While our data and models did not afford us the opportunity to determine the mechanisms by which seasonal fertility operates in the countries studied, our study did allow us to confirm that seasonal fertility exists in the absence of high-amplitude seasonal stressors such as reduced food availability and increased physical work. In countries where seasonal stressors still impact most of the population, seasonal fertility and seasonal sexual activity may combine to drive very strong birth seasonality pulses. This could occur, for instance, in seasonally migrant agricultural communities where household composition and behavioral activities are strongly tied to the seasonal environment, allowing for an alignment between physiology (fertility) and social activities (including sex). Globally, there are some countries (e.g., in Sub-Saharan Africa) with higher amplitude birth seasonality than those included in our study. It is important to understand how seasonal fertility and seasonal sexual activity operate in countries with these stronger seasonal rhythms.

Studies in Western countries have investigated if pregnancy planning may explain birth seasonality^8–10,31^. These studies showed that couples have a higher desire for births in the Spring or the Summer. In the US^31^, Denmark^31^, Sweden^10^, and, to some extent, in France^8^, pregnancy planning may play a role in shaping birth seasonality. However, in the US, as much as 49% of pregnancies may be unintended^32^, so we did not expect seasonal pregnancy planning to account for birth seasonality overall. Couples can implement seasonal pregnancy planning by interrupting contraception and/or increasing the frequency of unprotected sex at specific periods of the year. Our data showed similar temporal patterns for protected and unprotected sexual activity, with one important exception: a higher level of unprotected sex between Christmas and New Year, which explained elevated September births in most study locations. Therefore, unprotected sex had a subtle effect on birth seasonality. Still, our data suggest pregnancy planning did not play a significant role in shaping the birth curve, which we found to be shaped by seasonal fertility.

Various reproductive mechanisms could be driving seasonal fertility, including male and/or female fertility at conception and/or induced or spontaneous pregnancy loss. We show here that seasonal reproductive biology is the dominant underlying driver of birth seasonality and acts in combination with seasonal sexual activity to give the nuanced population-specific shape of birth curves. Our results raise the question of whether the external seasonal environment directly alters fertility or acts as a synchronizing factor to entrain an endogenous biological clock. Specifically, such endogenous circannual clocks that remain rhythmic under constant conditions have been known to exist in other mammals and contribute to seasonal physiology, including reproduction, hibernation, molting, and migration^33–35^.

Our study indicates peak fertility is centered around the winter solstice in both hemispheres. From the perspective of seasonal breeding biology, this would mean humans are short-day breeders (i.e., the human “breeding season” in which fertility peaks during the short days of winter). Initiating pregnancy during winter is in keeping with observations from other mammals with long gestation periods (e.g., sheep and deer) because gestation during the harsh winter months allows for births to be aligned with optimal food conditions and favorable environmental conditions in spring, summer, and early autumn. Our results are in line with other recent studies of fertility. Seasonal fecundability (i.e., fertility at conception) peaking in fall/winter was also observed in a cohort study in the Northern Hemisphere^31^, and increased endometrial receptivity at the same period of the year was suggested in an earlier study^23^.

In the Northern Hemisphere, the seasonal fertility peak around the winter solstice coincides with the dominant holiday season of New Year/Christmas when sexual activity is elevated. By contrast, in Brazil, in the Southern Hemisphere, the New Year/Christmas holiday is out-of-phase with the seasonal fertility peak. The inclusion of data from both the Northern and Southern Hemisphere is a key strength of our study, given that cultural holidays have emerged in seasonal environments. Therefore, including countries with differing seasonal cycles (e.g., temperature seasons vs. tropical rainy/dry seasons) allows us to reveal seasonal features of human biology that may otherwise be masked by cultural phenomena.

To our knowledge, this is the most in-depth study of sexual activity by way of the depth of variables relating to sexual activity, temporal resolution, geographical coverage, and sample size (over 500,000 app users). We found that sexual activity is strongly structured around holidays, weekends, and, in some locations, summer months. As expected, elevated sexual activity was generally associated with time off work/school. In addition, romantic celebrations, such as Valentine’s Day and Dia dos Namorados (i.e., Brazilian Valentine’s Day), expectedly have increased sexual activity. This is in agreement with sexual and reproductive health studies reporting an increase in sexual activity occurring around Christmas and other holidays and events^5,6^.

One limitation of our sexual activity data is that it originates from a self-selected, and therefore potentially biased, population of app users. However, it is important to note that the Clue app is used by a wide diversity of users, regardless of age, birth control use, health condition, or fertility status. The app is not targeted at sub-fertile individuals who may have different sexual activity patterns than the general population. When stratifying users by age or birth control use, we only observed slight differences in sexual activity. For example, in the US, older individuals reported more sex on Thanksgiving than younger ones. We speculate that older individuals may be more likely to be in relationships where they celebrate Thanksgiving with their sexual partners. In comparison, younger individuals may celebrate it with their families and not with their sexual partners.

Another limitation of our study was that our sexual activity data only covered two years. Although we observed consistency among those two years, we would have been able to quantify the uncertainty around our estimates of holiday sexual activity with more data. An interesting extension of this work would be to not only explore the interaction between day-of-week and holiday (for instance, if Christmas and the New Year were on weekends *vs*. midweek), but also include countries with different national holidays and explore the effect of holidays outside of the Judeo-Christian calendar, such as Jewish, Muslim, and Hindu holidays.

Altogether, our results show that although humans can reproduce year-round, there is seasonal structuring to our reproductive physiology. These results align with early findings of seasonal variation in reproductive hormones; specifically, increased estradiol in the winter^36^, which was recently confirmed by a large-scale study^37^ that revealed a winter-spring peak in estradiol and testosterone and a summertime peak in luteinizing hormone (LH) and prolactin ^37^. Humans also have seasonal cycles in hormones involved in thermoregulation, metabolism, stress adaptation, and growth^37^, as well as seasonal changes in the immune system^38,39^. While these changes might be subtle at the individual level, they may manifest as observable population-level phenomena, such as birth seasonality, which has clinical and public health implications. Times of elevated fertility may open a window of opportunity for reproduction, and individuals with lower fertility may benefit from synchronizing with their seasonal cycle for planning pregnancy. In contrast, the high fertility season is a high-risk period for individuals desiring or needing to avoid pregnancy. These risks and opportunities need to be recognized by the public health community to improve sexual and reproductive health interventions.

## Data Availability

Birth data, aggregated time series and model parameters are available on https://github.com/lasy/Seasonality-Public-Repo.

## Methods

We tested three hypotheses on birth seasonality using mathematical models of birth rates and combining sexual activity data collected from n = 535,154 individuals, representing 180 million days of active tracking data, with official birth record data in six locations. The six locations were selected based on the availability of data, such that they represent a geographically and culturally diverse set of countries/regions. Specifically, our population-level analyses were done for the UK and France at the country level and in two areas of the US and Brazil due to the geographic size and large population of the latter two countries. In the US, we used data from California, which is a Western state, and from the Northeast Census Region, which includes nine states on the Northeast Coast: Connecticut, Maine, Massachusetts, New Hampshire, Rhode Island, Vermont, New Jersey, New York, and Pennsylvania. In Brazil, we used data from the Central-West Region– which includes the states: Goiás, Mato Grosso, Mato Grosso do Sul, and Distrito Federal (which is the capital Brasilia) –and the Northeast Region, which is on the Northern Atlantic Coast and includes the states: Alagoas, Bahia, Ceará, Maranhão, Paraíba, Pernambuco, Piauí, Rio Grande do Norte, and Sergipe. Overall, our dataset includes these six geographic locations: the UK, France, Central-West Brazil, Northeast Brazil, California, and the Northeastern US (Fig 1a).

### Sexual activity data

Sexual activity data was collected from Clue, a women’s health mobile phone app (Fig 1b, main text). Our study was determined to be exempt from our institutional IRB because data were de-identified, and users were informed that their data might be shared for scientific research with the option to opt-out while still using the app. Since the app is specifically built for menstrual cycle tracking, we assume all users are female or menstruating individuals. The Clue app is developed and marketed to be used by a larger diversity of individuals, regardless of age, birth control methods, reproductive objectives (trying or avoiding conception), reproductive health status, gender, and culture. Importantly, the app is not specifically targeted to sub-fertile individuals. De-identified individual-level daily records were obtained from n = 535,154 Clue users. The number of users per location is: UK (n = 133,387), France (n = 126,634), Central-West Brazil (n = 160,896), Northeast Brazil (n = 23,127), California (n = 39,330) and the Northeastern US (n = 51,780). Clue users could prospectively log their menstrual bleeding, symptoms (e.g., headache, breast tenderness, etc.), and sexual activity each day using the app. In addition, for each user, Clue provided us with the user’s age and with their declared birth control method. However, we were not provided with reports of pregnancy or miscarriage.

As with all tracking apps, there was variation in the fidelity with which users tracked. Overall, we obtained n = 55,110,476 logged daily records from July 2017 to July 2019, spanning a period of 180 million days of active tracking. It is important to note that individuals may be actively tracking but will not necessarily have something to log each day of their cycle. Due to changes in tracking fidelity, we used a rolling window to identify periods of active tracking for each individual (refer to Suppl. Mat. Section 2.4 for more details). Sexual activity was reported as three types: protected, unprotected, or withdrawal sex.

For each geographic location and sex type, aggregated time-series of population-level daily sexual activity were constructed. These time series represented the daily sex rate in the population as defined by the fraction of active users reporting a specific type of sex. Because these data are not ideal for estimating the absolute amount of sexual activity in a population (*i*.*e*., due to biases in app user demographics and variation in tracking fidelity), we computed the relative changes in sexual activity by dividing the sex counts by the number of users that opened the app on each day (Fig 1c). This allowed us to focus on variation around the mean, particularly variation tied to day-of-week, holidays, and seasons.

To infer how sexual activity is impacted by holidays, weekends, and seasons, we fitted a statistical model to our data to measure relative sexual activity. This model allowed us to predict sexual activity for any location and any given calendar day in the year, based on the day-of-week, month, and proximity to a local holiday (Fig 2). Due to the large sample size, we were able to fit this statistical model for all combinations of (i) age, (ii) birth control method (barrier methods vs. hormonal or IUD), and (iii) protected vs. unprotected sex (see section 4.3.2 of the Supplementary Material). Since the sexual activity was similar among groupings, results in the main text and figures are based on unprotected sex reported by users of any age and birth control category.

Our model was structured as follows: sex[*i, j*] = *α*_*i*_ h[*i*] + *β*_*j*_ wdm[*j*] where each 365 days of the year is defined by a specific combination of *i* and *j*. The first term represents the contribution of a particular holiday to sexual activity and the second term represents the contribution of the day-of-week and month. Each holiday has its own coefficient because not all holidays lead to the same increase or decrease in sexual activity. Based on official national holiday calendars, all holidays included in our models have a day off work/school, except for Valentine’s Day and the Dia dos Namorados (*i*.*e*., Brazilian Valentine’s Day). For holidays with a day off, we estimated a fixed amount of sexual activity in the first term while the second term is set to its intercept value because we assumed the weekday and month effect was negligible when people are given a day off. Whereas, for Valentine’s and Dia dos Namorados, relative sexual activity was estimated as the sum of the holiday contribution combined with the weekday and month effect. This accounted for the fact that these holidays fall on different days of the week each year and don’t lead to a day off. In addition, we assumed there were three days off before and after Christmas and New Year and estimated a fixed amount of sexual activity on those days. Refer to section 4.3.2 of the Supplementary Materials for the holiday description (*i*.*e*., holiday categorization based on whether each holiday results in a day off of work/school).

To validate that the holiday and weekend sexual activity patterns were not an artifact relating to more app use on the holidays and weekends, we tested if biological variables reported by Clue users displayed the same patterns as sex. This is because we would not expect biological variables such as vaginal bleeding to be more prevalent on holidays and weekends. Thus, a weekly or holiday pattern in vaginal bleeding would reveal reporting bias due to app use. We did not observe holiday and weekend patterns in our biological variables and concluded that holiday and weekend patterns of sexual activity were reflective of real-world changes and not an artifact of reporting on the app.

### Birth Data

Time series of monthly live births were obtained for each geographical area. The US data were obtained from the CDC Wonder database and span 2007 to 2018. Brazilian birth data were obtained from DATASUS and span 2000-2016. UK data are from the Office of National Statistics and span 2000-2018, while data from France were retrieved from the INSEE database for years 2005-2019. Due to variation in the number of days per calendar month, the time series were standardized to represent a typical 30-day month. We did this to ensure that seasonality in the time series was not an artifact of variation in days per month.

### Mathematical Models

To test if birth seasonality was driven by seasonal changes in sexual activity, fertility, or both, we translated each of our three birth seasonality hypotheses into a deterministic population-level mathematical model of conception and birth (Fig 3a). The hypotheses were: Hypothesis A: birth seasonality is solely driven by seasonal changes in sexual activity, Hypothesis B: birth seasonality is solely driven by seasonal fertility, and Hypothesis C: birth seasonality is driven by a combination of seasonal sexual activity and seasonal fertility. Daily conceptions were modeled as the product of daily sexual activity and daily fertility in each model. Daily births were then predicted by shifting daily conceptions by approximately nine months into the future using empirical distributions of gestation periods for each country (Suppl. Fig. 17)^25-27^. Since the time series of births were monthly totals, we aggregated simulated daily births to monthly. Parameters of each model were estimated by minimizing the sum of squared errors between simulations and data. For each model, we calculated the likelihood and the AIC, which includes a penalty for additional parameters. We then discriminated among models using AIC.

Model A assumed variation in sexual activity entered as a time-varying variable in the model and constant fertility (Fig 3a). The time-series of daily sexual activity used in Model A was the output from our statistical model fitted to the Clue app data. Model B assumed constant sexual activity and seasonal variation in fertility expressed as a sinusoidal function with a one-year period (Fig 3a). The phase and amplitude of this function were estimated for each geographical area. Model C assumed variation in both sexual activity and fertility (Fig 3a). For Model C, seasonal sexual activity was entered as the time-series of daily sexual activity output from our statistical model (as for model A). We added a scaling factor to modulate the amplitude of sexual activity. This scaling factor, along with the phase and amplitude of seasonal fertility, were estimated independently for each geographical location. Lastly, the mean daily conception rate was calculated based on the long-term trend in the birth time series.

## Acknowledgments

The authors thank V. Teixeira and J. Calixto for helping extract the Brazilian birth data from DATASUS and Dr. Vitzthum for helpful feedback on the manuscript.

## Authors’ contributions

L.S., M.M. contributed to the conceptualization and methodology. L.S., A.S., C.R.M., D.S., and M.M. contributed to the data acquisition and curation. L.S. contributed to the software and visualizations. L.S., M.M., P.S., D.S., C.R.M, and S.H. contributed to the writing of the manuscript.

## Data and materials availability

code, birth data, aggregated time series, and models are available on https://github.com/lasy/Seasonality-Public-Repo.

